# Platelet Function Test-Guided Antiplatelet Therapy Reduces Recurrent Stroke in Atherothrombotic and Lacunar Infarction

**DOI:** 10.64898/2026.04.06.26350274

**Authors:** Shintaro Nakayasu, Hideki Hayashi, Ryohei Goda, Masaki Maeda, Nao Yoshioka, Shinya Kobayashi, Eiji Ogino, Fumihiko Horikawa, Nozomu Murai

**Affiliations:** Uji-Tokushukai Medical Center

## Abstract

**Background and Purpose:** Antiplatelet resistance is a recognized risk factor for recurrent ischemic stroke, yet evidence supporting platelet function test–guided antiplatelet therapy modification in stroke prevention remains limited. We investigated whether VerifyNow-guided antiplatelet therapy modification reduces recurrent ischemic stroke in patients with atherothrombotic or lacunar infarction.

**Methods:** This retrospective observational study enrolled consecutive patients with atherothrombotic or lacunar infarction at a single center (April 2023–March 2025). Of 302 patients, 243 were analyzed: 122 in the modified group, whose antiplatelet agent was selected based on VerifyNow Aspirin Reaction Units and P2Y12 Reaction Units, and 121 in the unmodified group, whose agent was empirically selected. The mean follow-up period was 1.62 ± 0.61 years. In the modified group, when both aspirin and clopidogrel showed inadequate inhibition, prasugrel or cilostazol was selected. The primary endpoint was recurrent ischemic stroke; the secondary endpoint was intracranial hemorrhage. Cox proportional hazards models with inverse probability weighting were used to adjust for confounders.

**Results:** Recurrent ischemic stroke occurred in 1 patient (0.8%) in the modified group versus 8 (6.6%) in the unmodified group (log-rank P=0.018). After adjustment, the modified group had a significantly lower risk of recurrent stroke (HR, 0.10; 95% CI, 0.012–0.84; P=0.033). Intracranial hemorrhage occurred in 0 (0%) and 1 (0.8%) patients, respectively.

**Conclusions:** In Japanese patients with atherothrombotic or lacunar infarction, VerifyNow-guided antiplatelet therapy modification was associated with a significantly lower incidence of recurrent ischemic stroke without increased hemorrhagic risk. Given the single-center retrospective design and small sample size, validation in a multicenter randomized controlled trial is warranted.

## Introduction

Ischemic stroke is one of the most common causes of death and long-term disability worldwide [1]. Cerebral infarction is frequently recurrent, occurring in approximately 11% of patients within one year and 39.2% within ten years [2]. Antiplatelet therapy is recommended in guidelines for preventing cerebral infarction recurrence, and both aspirin and clopidogrel are frequently prescribed antiplatelet agents. However, there is interindividual variability in platelet responsiveness to both aspirin and clopidogrel. The prevalence of non-response to aspirin and clopidogrel is reported to be 23% for aspirin and 27% for clopidogrel [3]. Previous studies suggest that both aspirin resistance and clopidogrel resistance are risk factors for stroke and adverse vascular events [4, 5]. There are reports indicating that individualized antiplatelet therapy based on platelet responsiveness testing is beneficial for preventing ischemic complications after endovascular treatment in cerebral regions [6]. Information on the efficacy of changing antiplatelet agents based on VerifyNow results is limited.

Therefore, the objective of this study was to investigate clinical outcomes associated with antiplatelet therapy modification guided by platelet function testing in patients with ischemic stroke. The hypothesis was that antiplatelet therapy modified based on platelet function testing would reduce the incidence of recurrent stroke without increasing bleeding risk.

## Methods

### Study Design and Participants

This retrospective observational study enrolled patients diagnosed with atherothrombotic cerebral infarction or lacunar infarction who underwent inpatient treatment at our hospital. Consecutive participants were sequentially enrolled at our hospital from April 1, 2023, to March 31, 2025.

Selection criteria were as follows:

1. Presence of cerebral infarction confirmed by magnetic resonance imaging (MRI).
2. Undergoing computed tomography angiography (CTA) or magnetic resonance angiography (MRA) of the brain and carotid arteries, or color duplex ultrasound of the carotid arteries.
3. Patients who underwent routine electrocardiogram (ECG), 24-hour Holter ECG, and echocardiography to exclude cardioembolic stroke
4. Patients admitted to the ward who received treatment according to the 2021 Japan Stroke Guidelines for Acute Ischemic Stroke. [7]
5. During the screening period, a series of investigations including blood sampling were performed, and strokes were classified into five subtypes according to the criteria of the Trial of Org 10172 in Acute Stroke Therapy (TOAST) [8]. Only the atherothrombotic and small vessel disease subtypes were included.

Adherence was determined through interviews with patients and caregivers. Exclusion criteria were as follows:

1. Requirement for oral anticoagulants due to clinically significant arrhythmias (including atrial fibrillation) or deep vein thrombosis
2. Cases deemed to require continued dual antiplatelet therapy in the chronic phase
3. Difficulty in follow-up outside the medical care area
4. Death at discharge or best supportive care policy
5. Poor medication adherence
6. Allergy to aspirin or clopidogrel
7. Inability to perform detailed imaging evaluation or obtain sufficient medical history

Participants provided written informed consent for participation.

### Platelet Reactivity Assessment

Platelet function was measured using the VerifyNow Aspirin and the VerifyNow P2Y12 (Accumetrics, Inc, San Diego, CA).

### Standard Operating Procedure for Antiplatelet Medication Adjustment

All patients received either the standard combination dose of aspirin 100 mg and clopidogrel 75 mg once daily, or the standard single-agent dose of aspirin 100 mg or clopidogrel 75 mg once daily, starting at least 7 days prior to undergoing the VerifyNow Aspirin and VerifyNow P2Y12. Additionally, except for cilostazol, a loading dose was administered at the initial dose for first-time antiplatelet therapy (aspirin 300 mg, clopidogrel 300 mg, or prasugrel 20 mg). Participants in the modification group underwent VerifyNow platelet function testing at least 7 days after starting antiplatelet therapy. Based on previous research findings, dual antiplatelet therapy was continued for at least three weeks following the onset of cerebral infarction [9]. Subsequently, in the modified group, the antiplatelet agent deemed more effective based on VerifyNow platelet function test results was selected. In cases where both aspirin and clopidogrel were considered ineffective, prasugrel or cilostazol was selected. In the non-modified group, as in the modified group, dual antiplatelet therapy was administered for at least 3 weeks. Subsequently, based on the attending physician’s judgment, a single antiplatelet agent was selected and continued.

### Outcome Assessment

Recurrent ischemic stroke was defined as the primary endpoint. Intracranial hemorrhage was defined as a secondary endpoint. Outcomes were determined by researchers reviewing medical records.

### Data Collection

The following variables were collected and analyzed: patient demographics, including age and sex; and clinical characteristics, including history of hypertension, diabetes, prior stroke, coronary artery disease, and smoking. Data were obtained from MRI, MRA, and carotid Doppler ultrasound to assess intracranial and extracranial arterial stenosis or cerebral small vessel disease. Angiographic cerebral artery stenosis was primarily assessed from MRA, with ≥50% stenosis classified as severe. Concomitant medications, including antihypertensives(angiotensin-converting enzyme (ACE) inhibitor/angiotensin II receptor blockers (ARB), calcium channel blocker (CCB)), statins, and proton pump inhibitors (PPI), were recorded. Recurrent ischemic stroke and intracranial hemorrhage were recorded during follow-up.

### Statistical Analysis

Demographic and clinical variables were analyzed using descriptive statistics. Continuous variables were presented as mean ± standard deviation. Comparisons between groups for continuous variables were performed using independent samples t-tests. Categorical variables were presented as frequencies and analyzed using Fisher’s exact test. Statistical significance was defined as a two-sided P value < 0.05. Survival analysis was used to evaluate the difference between the modified and unmodified groups for the primary endpoint, estimating hazard ratios (HR), Kaplan-Meier plots, and log-rank test P values. Multivariable analysis of survival time was performed using Cox proportional hazards models with inverse probability of treatment weighting (IPTW) for adjustment of confounding factors (age, hypertension, diabetes, coronary heart disease, smoking, previous stroke, proton pump inhibitor, intracranial artery stenosis, extracranial artery stenosis, antiplatelet therapy). All statistical analyses were performed using EZR version 1.70. EZR is statistical software that extends the functionality of R and R Commander, and is freely distributed on the website of Saitama Medical Center, Jichi Medical University.

### Ethical review

This study was approved by the Ethics Committee of Uji-Tokushukai Medical center (approval number: 2025-23)

## Results

A total of 302 participants were enrolled in this study. Among them, 129 patients had an atherothrombotic brain infarction and 173 patients had a lacunar infarction. A total of 59 participants were excluded. Of the 59 exclusions, 20 were due to anticoagulant use, 12 to dual antiplatelet therapy, 6 to being outside the medical service area, 3 to poor adherence, 4 to lack of MRI data or missing data, and 14 to death or best supportive care. Details of patient selection are shown in Figure 1. Baseline characteristics of the modified antiplatelet therapy group and the unmodified group are shown in Table 1. The mean follow-up period was 1.62 ± 0.61 years, with 1.63 ± 0.59 years in the modified group and 1.60 ± 0.63 years in the unmodified group. Standardized mean differences exceeded 0.1 for hypertension, CCB, and PPI, indicating imbalance between groups. The primary endpoint occurred in 1 case (0.8%) in the modified group and 8 cases (6.6%) in the unmodified group. The secondary endpoint occurred in 0 cases (0%) in the modified group and 1 case (0.8%) in the unmodified group. Table 2 shows the antiplatelet agents used in the modified and unmodified groups. Table 3 also shows the ARU values and PRU values that served as the basis for selecting antiplatelet agents in the modified group for each antiplatelet drug. Clopidogrel was selected more frequently in the modified group, while cilostazol was selected more frequently in the unmodified group. Kaplan-Meier analysis showed a significantly lower incidence of recurrent ischemic stroke in the modified group compared with the unmodified group (log-rank test, P=0.0176, Figure 2). Furthermore, Cox proportional hazards regression analysis adjusted for confounders using inverse probability weighting demonstrated a significant difference in the primary endpoint between the modified and unmodified groups (HR 0.10, 95% CI 0.012–0.84, P=0.033, Table 4). Subgroup analysis in the group with severe intracranial arterial stenosis showed a higher incidence of recurrent cerebral infarction. However, due to the small number of cases and events, the statistical power was low, and no significant difference in the primary endpoint was observed after adjustment for antiplatelet therapy (modified group, 1/34, 2.9%; unmodified group, 4/26, 15.4%, P=0.156, Table 5). The results suggest that antiplatelet drug modification according to VerifyNow platelet function testing may reduce the incidence of recurrent stroke.

**Fig. 1:**
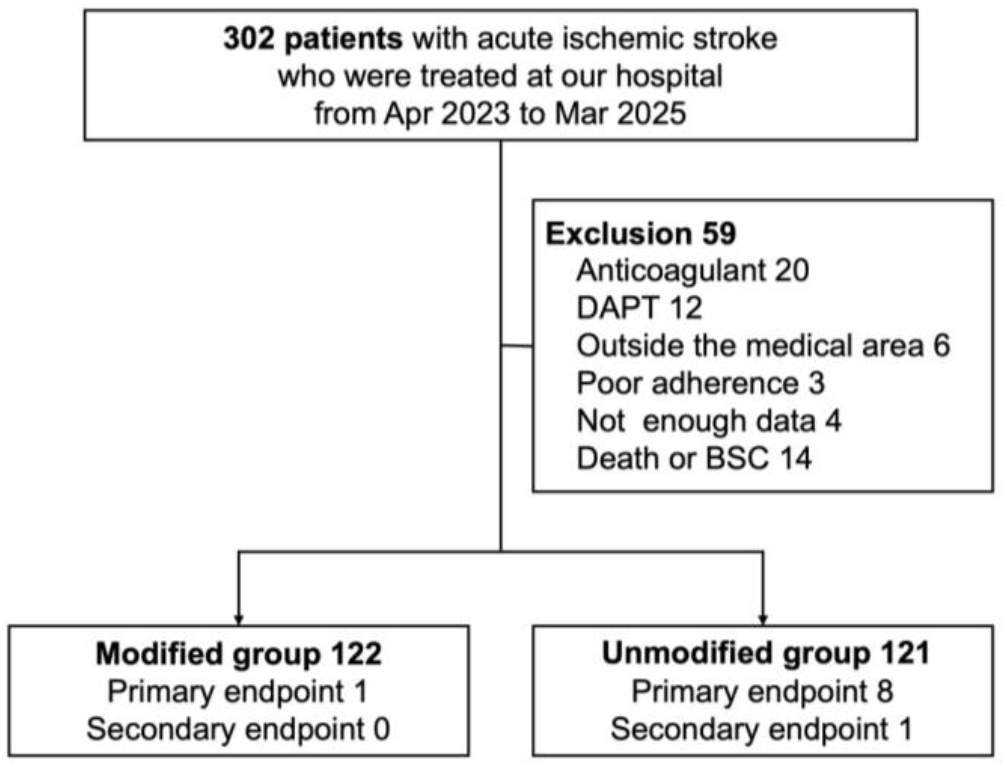
Details of patient selection

**Table 1:** Baseline characteristics of the modified antiplatelet therapy group and the unmodified group.

|  | Modified group (n=122) | Unmodified group (n=121) | SMD |
| --- | --- | --- | --- |
| Age | 73.69±10.39 | 74.12±12.61 | 0.037 |
| Male | 77 (63.1) | 72 (59.5) | 0.074 |
| Hypertension | 99 (81.1) | 83 (68.6) | 0.292 |
| Diabetes | 33 (27.0) | 34 (28.1) | 0.023 |
| Coronary heart disease | 17 (13.9) | 10 (8.3) | 0.181 |
| Smoking | 61 (50.0) | 65 (53.7) | 0.074 |
| Previous stroke | 23 (18.9) | 18 (14.9) | 0.106 |
| CCB | 81 (66.4) | 62 (51.2) | 0.312 |
| ACE/ARB | 64 (52.5) | 64(52.9) | 0.009 |
| Statin | 65 (53.3) | 70 (57.9) | 0.092 |
| PPI | 117 (95.9) | 106 (87.6) | 0.305 |
| Intracranial artery stenosis | 76 (62.3) | 79 (65.3) | 0.062 |
| Severe cereberal arterial stenosis | 34 (27.9) | 26 (21.5) | 0.148 |
| Extracranial arterial stenosis | 32 (26.2) | 22 (18.2) | 0.195 |
| Primary endpoint | 1 (0.8) | 8 (6.6) | 0.31 |
| Secondary endpoint | 0 (0) | 1 (0.8) | 0.129 |

**Table 2:**
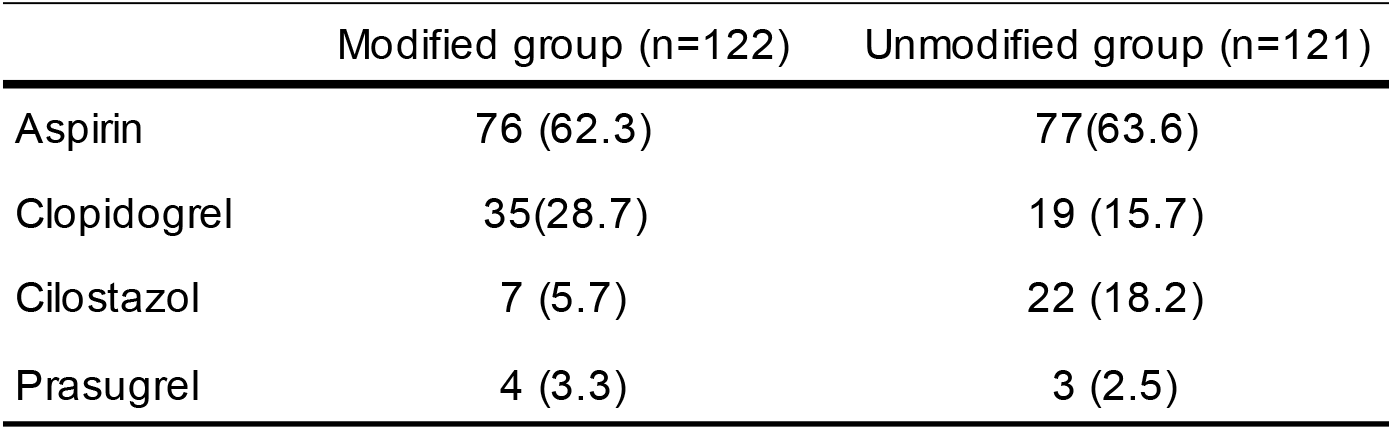
Antiplatelet agents used in the modified and unmodified groups.

**Table 3:** ARU and PRU values in the modified group stratified by selected antiplatelet agent.

|  | ARU | PRU |
| --- | --- | --- |
| Aspirin (n=76) | 414.24 (±62.82) | 209.03 (±68.58) |
| Clopidogrel (n=35) | 465.31 (±79.15) | 171.49 (±45.46) |
| Cilostazol (n=7) | 563.29 (±37.73) | 228.43 (±33.93) |
| Prasugrel (n=4) | 446.00 (±106.95) | 267.25 (±191.89) |

**Fig. 2:**
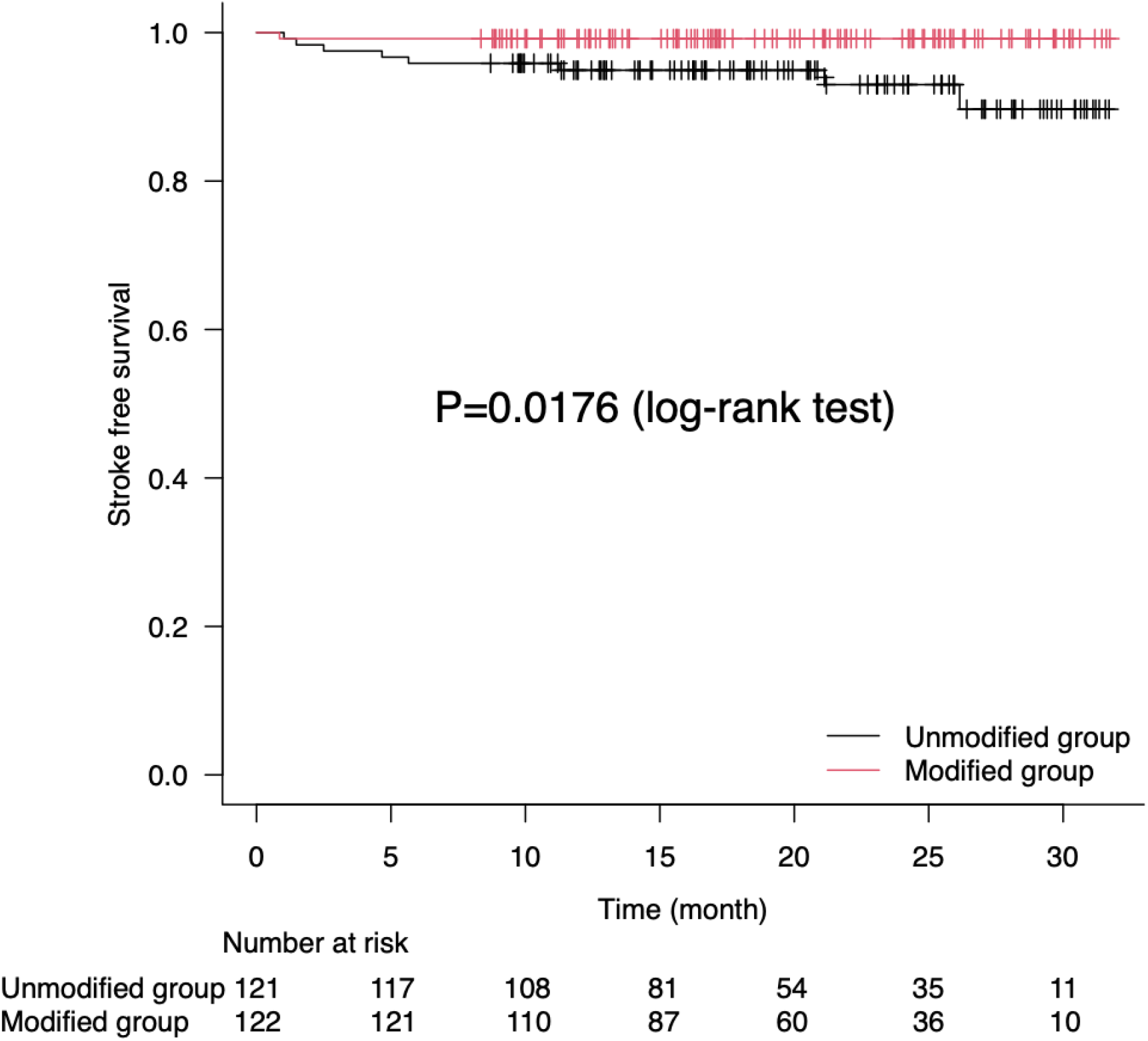
Kaplan-Meier analysis for primary outcome comparison between the modified group and unmodified group

**Table 4:**
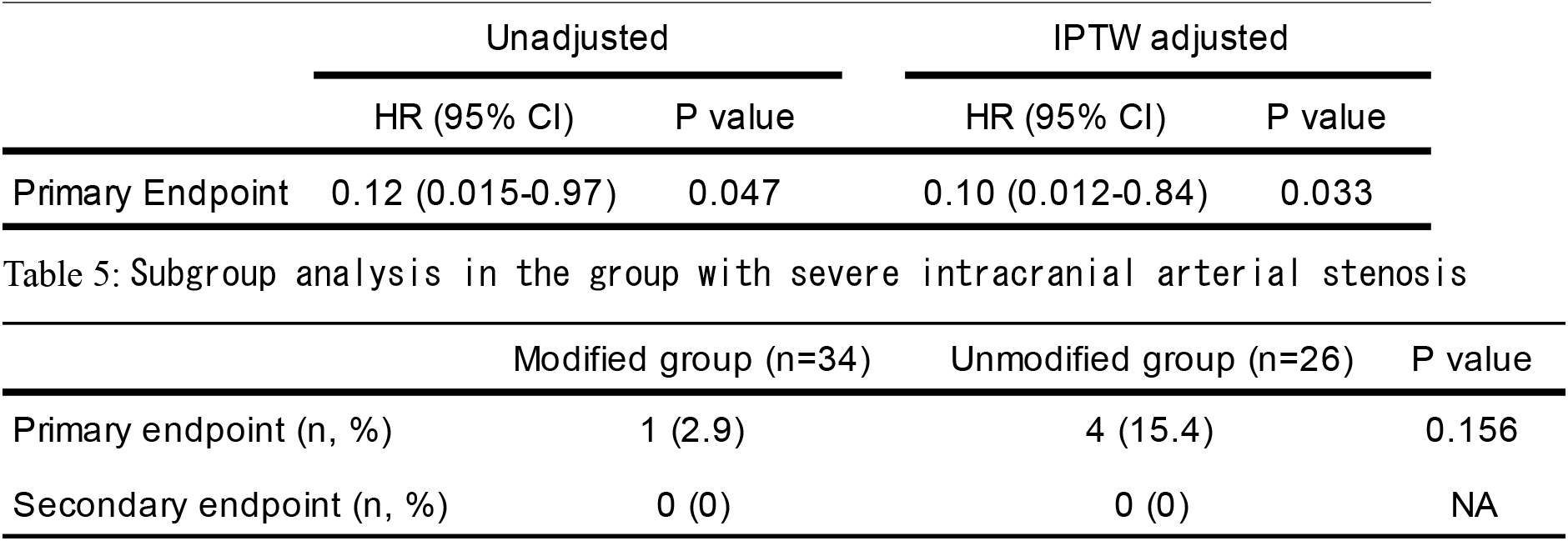
Cox proportional hazards regression analysis with inverse probability weighting for the primary endpoint.

## Discussion

### Summary of Research Findings

Our results indicate that modifying antiplatelet therapy based on VerifyNow platelet function testing, after adjusting for severe intracranial and extracranial arterial stenosis, may reduce the incidence of recurrent ischemic stroke without increasing bleeding risk. This suggests that selecting appropriate antiplatelet therapy may achieve long-term prevention of stroke recurrence. Outcome rates remained significantly different between the modified and unmodified groups even after adjusting for confounders, indicating that recurrent ischemic stroke induced by inadequate platelet inhibition may be mitigated by appropriate antiplatelet modification.

### Technical Features and Convenience of VerifyNow Platelet Function Testing

VerifyNow platelet function testing enables rapid and simple monitoring of platelet function using whole blood via optical turbidimetric aggregation. It is characterized by its ability to quantify drug responsiveness using metrics such as P2Y12 reaction units (PRU) and aspirin reaction units (ARU) [10]. In clinical practice, it has gained attention as a powerful tool, particularly in the PCI field, for assessing thrombosis risk after stent placement [11]. Its usefulness has also been suggested in intracranial vascular stenosis and endovascular cerebrovascular therapy [12,13].

### Heterogeneity of Pathogenesis in Cerebrovascular Disorders and Challenges in Applying VerifyNow Platelet Function Test Data

The pathogenesis of TIA and stroke is highly heterogeneous, and caution should be exercised when directly applying in vitro results. In vitro tests may not adequately replicate in vivo conditions such as shear stress in blood flow. It has been pointed out that these tests may not fully replicate aspects of in vivo thrombus formation, such as the simultaneous action of multiple agonists, thrombin generation, interactions with the vascular wall, and evaluation of the fibrinolytic system [14]. Consequently, major RCTs such as the GRAVITAS and ARCTIC trials have all failed to demonstrate that modified antiplatelet therapy guided by the VerifyNow platelet function test improves clinical outcomes [15,16].

### Inconsistency in Cutoff Values and Ambiguity in Standards

Cutoff values for PRU and ARU in VerifyNow platelet function testing vary across studies, and no globally unified standard currently exists. While PRU > 208 or 230 is commonly used to define HPR (high platelet reactivity), some reports suggest PRU ≥ 240 is useful for predicting prognosis at one month [17]. For Asian patients, considering bleeding risk and the context of the “East Asian paradox,” a distinct cutoff value such as PRU 190 has been proposed to improve prognosis [5].

### Reproducibility and Temporal Variability of Test Results

VerifyNow platelet function testing, particularly regarding reactivity to clopidogrel (PRU values), has been noted as having significant issues with reproducibility between assays and temporal variability of test results [18]. Data indicate that 38.2% of patients experienced at least one change in responder status between 1 day and 6 months post-PCI. This tendency is particularly pronounced with clopidogrel, where a weakening of antiplatelet effect and emergence of resistance have been observed during the maintenance phase compared to the initial treatment period [19].

### Comparison with Other Research Findings

Previous studies have evaluated VerifyNow-guided therapy for secondary stroke prevention, but the major difference lies in the criteria for selecting antiplatelet agents. In all prior studies, either aspirin or clopidogrel alone was used as the first-line antiplatelet agent, and VerifyNow platelet function testing was performed to modify the antiplatelet agent in cases of non-response [4,5]. In this study, both VerifyNow platelet function tests for aspirin and clopidogrel were performed in all patients in the modification group, and the antiplatelet agent with the more favorable value was selected. Furthermore, in cases where both were judged ineffective, either cilostazol or prasugrel was selected. This antiplatelet selection strategy, considering both aspirin resistance and clopidogrel resistance, may have been useful for preventing recurrence.

Furthermore, all patients in this study were Japanese. It is known that Asians, particularly those with a high prevalence of loss-of-function alleles in CYP2C19, exhibit a high frequency of clopidogrel resistance [20]. This suggests that our population may have been more likely to benefit from antiplatelet therapy modification.

Previous studies, after adopting severe vascular stenosis as a covariate, demonstrated a significant difference in cerebral infarction recurrence following VerifyNow-guided intervention. Past research has also reported an association between severe cerebral vascular stenosis and stroke recurrence [21,22]. Our study also observed a high rate of recurrent cerebral infarction in cases with severe intracranial vascular stenosis (Table 5). In the subgroup with severe intracranial arterial stenosis, although the analysis did not reach statistical significance in the primary endpoint because this study had low statistical power due to the small number of cases and events, modifying antiplatelet therapy may be more beneficial in cases with severe intracranial vascular stenosis considering its high recurrence rate.

### Limitations

Several important limitations should be considered. First, this study was a single-center retrospective observational study, which may limit the generalizability of the results. Second, the diverse changes in antiplatelet therapy used after platelet function testing were at the physician’s discretion. The clinical factors that led each physician to decide which treatment regimen to use after platelet function testing are unknown, making control of selection bias very difficult. Third, although careful analyses were performed to account for any differences between patients in the antiplatelet therapy modification group and the non-modification group, unknown confounding factors may have contributed to differences in clinical outcomes between the two groups. Fourth, intracranial vascular assessment in this study was primarily performed using MRA, which may be less accurate than assessments using CTA or DSA. Furthermore, this study had a relatively small sample size and an extremely low number of events. Therefore, our findings must be validated in a multicenter study with a larger sample size and a randomized controlled trial.

## Conclusions

In this retrospective observational study of Japanese patients with atherothrombotic or lacunar infarction, modification of antiplatelet therapy guided by VerifyNow platelet function testing was associated with a significantly lower incidence of recurrent ischemic stroke compared with empirically selected antiplatelet therapy, without an increase in intracranial hemorrhage. The comprehensive assessment of both aspirin and clopidogrel responsiveness, with alternative selection of prasugrel or cilostazol in cases of dual non-response, may represent a practical approach for individualized secondary stroke prevention, particularly in populations with a high prevalence of clopidogrel resistance. However, given the single-center retrospective design, small sample size, and low number of events, these findings should be interpreted with caution and warrant validation in a multicenter randomized controlled trial with a larger sample size.

## Data Availability

The data that support the findings of this study are available from the corresponding author upon reasonable request.

## Non-standard Abbreviations and Acronyms

ACE: angiotensin-converting enzyme
ARB: angiotensin II receptor blockers
CCB: calcium channel blocker
PPI: proton pump inhibitors
IPTW: inverse probability of treatment weighting

## Acknowledgments

none

